# Specificity of SARS-CoV-2 antibody-detection assays against S and N protein among pre-COVID-19 sera from patients with protozoan and helminth parasitic infections

**DOI:** 10.1101/2021.08.10.21261841

**Authors:** Cedric P Yansouni, Jesse Papenburg, Matthew P. Cheng, Rachel Corsini, Chelsea Caya, Fabio Vasquez Camargo, Luke B Harrison, Gerasimos Zaharatos, Philippe Büscher, Babacar Faye, Magatte Ndiaye, Greg Matlashewski, Momar Ndao

## Abstract

**Background:** We aimed to assess the specificity of SARS-CoV-2 antibody detection assays among people with known tissue-borne parasitic infections.

**Methods:** We tested three SARS-CoV-2 antibody-detection assays (cPass SARS-CoV-2 Neutralization Antibody Detection Kit, Abbott SARS-CoV-2 IgG assay, and STANDARD Q COVID-19 IgM/IgG Combo Rapid Test) among 559 pre-COVID-19 sera.

**Results:** The specificity of assays was 95-98% overall. However, lower specificity was observed among sera from patients with protozoan infections of the reticuloendothelial system, such as human African trypanosomiasis (Abbott Architect; 88% [95%CI 75-95]), visceral leishmaniasis (SD RDT IgG; 80% [95%CI 30-99]), and from patients with recent malaria from a holoendemic area of Senegal (ranging from 91% for Abbott Architect and SD RDT IgM to 98-99% for cPass and SD RDT IgG). For specimens from patients with evidence of past or present helminth infection overall, test specificity estimates were all ≥ 96%. Sera collected from patients clinically suspected of parasitic infections that tested negative for these infections yielded a specificity of 98-100%. The majority (>85%) of false-positive results were positive by only one assay.

**Conclusions:** The specificity of SARS-CoV-2 serological assays among sera from patients with tissue-borne parasitic infections was below the threshold required for decisions about individual patient care. Specificity is markedly increased by the use of confirmatory testing with a second assay. Finally, the SD RDT IgG proved similarly specific to laboratory-based assays and provides an option in low-resource settings when detection of anti-SARS-CoV-2 IgG is indicated.

## INTRODUCTION

Use cases for serological testing for *Severe acute respiratory syndrome-related coronavirus-2* (SARS-CoV-2) have been reviewed in detail (1, 2). Despite a rapid increase in the number and availability of serological assays detecting anti-SARS-CoV-2 antibodies, critical knowledge gaps remain regarding cross reactivity of assays with sera from patients with non-coronavirus infectious agents.

In tropical regions of the world, several infections that dominate the local epidemiology of acute fever syndromes may cause non-specific cross reactivity with a wide range of serological assays (3-7). The mechanisms underlying cross reactivity include infection of the reticuloendothelial system by several protozoans, with associated polyclonal B-lymphocyte proliferation (6, 7), and the broad diversity of antibodies elicited by various helminth infections (8). These infections include many Neglected Tropical Diseases (NTD) and Malaria, for which the combined global burden exceeds 1.6 billion cases annually, with 3.8 billion at risk (9, 10). Simultaneously, three of the four countries with the largest total number of deaths attributed to COVID-19 are currently Brazil, India, and Mexico (11), all of which suffer a high burden of NTDs or malaria. As a result of this overlapping incidence, the specificity of SARS-CoV-2 serological tests may be different in countries endemic for these infections compared to that in high-income countries.

We aimed to assess the specificity of SARS-CoV-2 antibody detection assays among people either with known tissue-borne parasitic infections or residing in areas where such infections are endemic. We tested three SARS-CoV-2 antibody-detection assays against either the S or the N protein, among a large collection of pre-COVID-19 sera from patients proven to have different tropical infectious diseases.

## METHODS

### Ethics

This work was approved by the Research Ethics Boards of the Research Institute of the McGill University Health Centre (RI-MUHC # 2021-7246).

### Source of specimens tested

Specimens were well-characterised sera collected prior to July 2019 from patients with active or recent malaria imported to Canada, with active or recent malaria in a hyperendemic area (Senegal), from clinical suspects for human African trypanosomiasis and for visceral leishmaniasis, from seropositives for Chagas disease, for *Strongyloides* sp., for *Schistosoma* sp., for filaria species, and for *Trichinella* sp., and from negative controls for whom tissue-borne parasitic infection was suspected but antibody testing was negative. The source and types of specimens are detailed in Table 1.

**Table 1.**
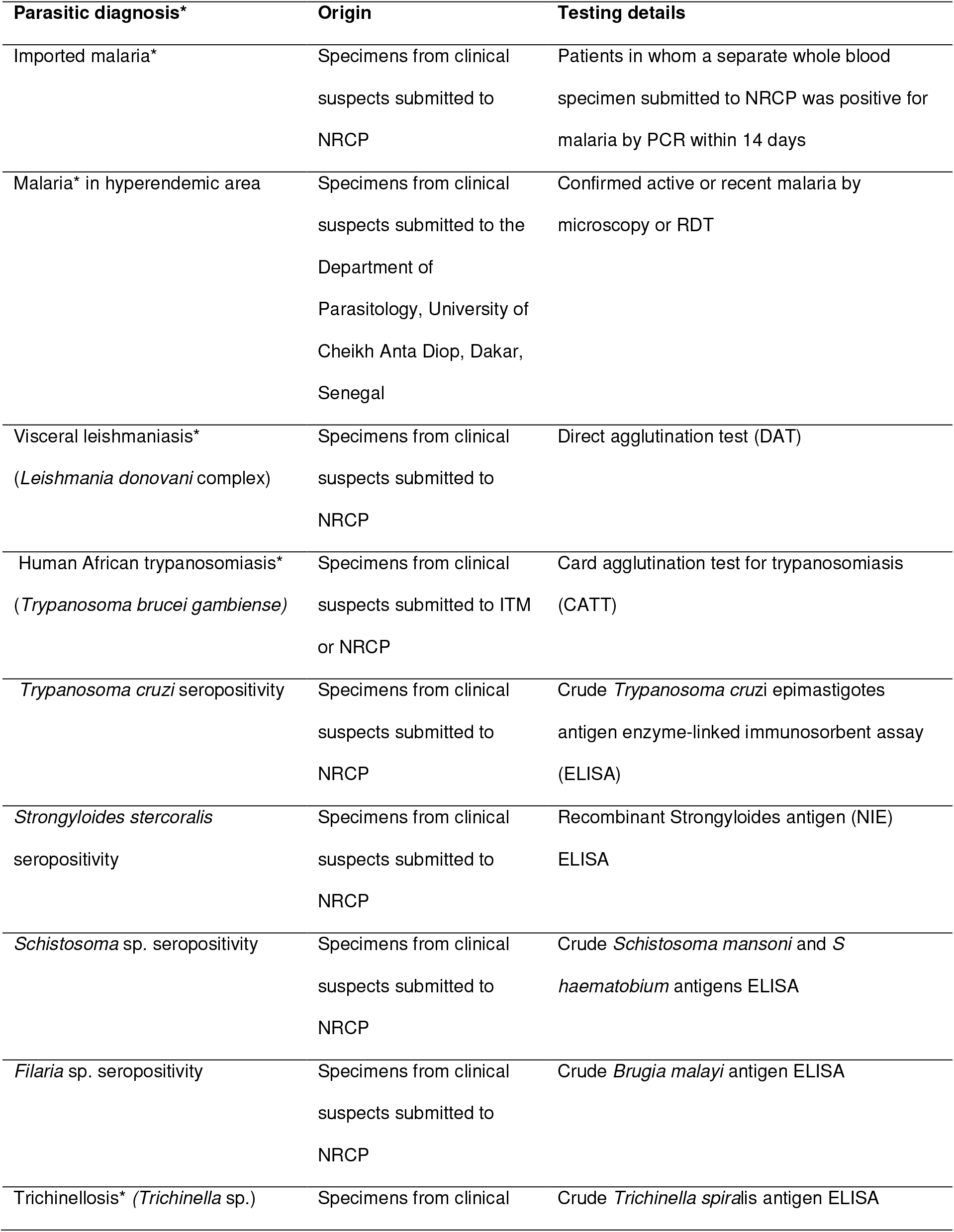
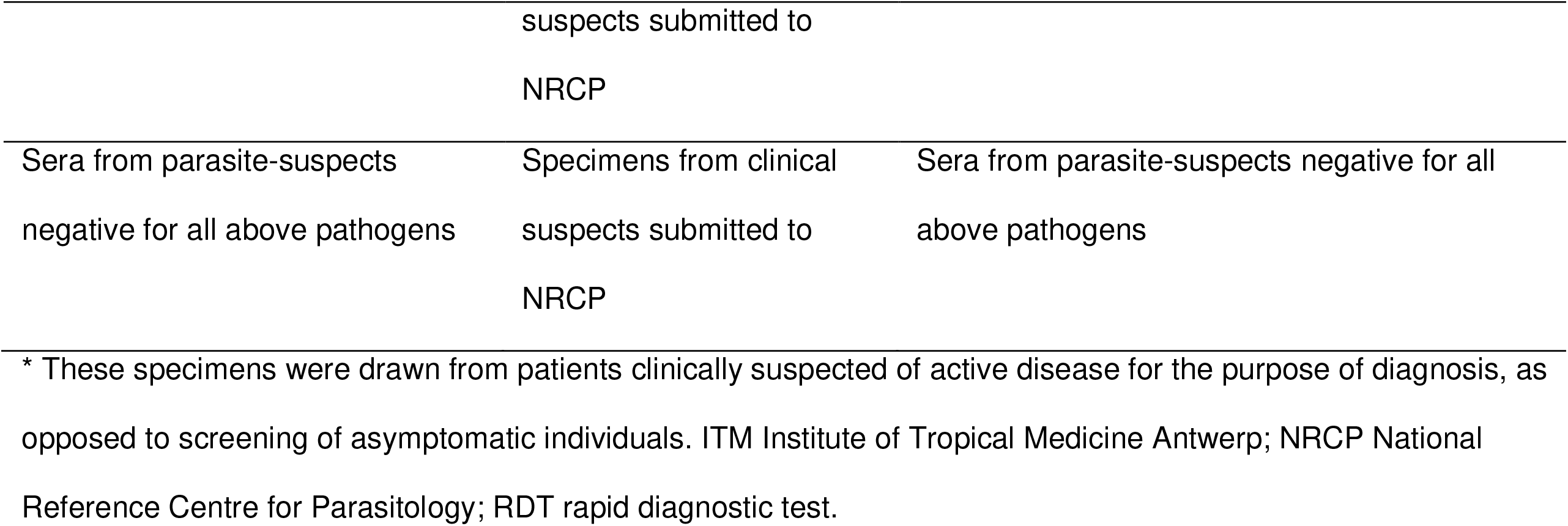
Origin of Pre-COVID-19 specimens.

### SARS-CoV-2 antibody testing

Three different SARS-CoV-2 antibody-detection assays were selected to assess the specificity of assays that detect different analytes, including anti-SARS-CoV-2 N-protein IgM, anti-SARS-CoV-2 N-protein IgG, and anti-Receptor Binding Domain (RBD) blocking antibodies of all immunoglobulin subclasses. We included an immunochromatographic rapid diagnostic test (RDT) that can be performed in low resource settings and available from a quality-assured manufacturer with an international presence to enhance the relevance of this evaluation to the low resource settings where NTDs and malaria are common.

### Culture-free neutralization antibody detection assay (cPass)

The cPass SARS-CoV-2 Neutralization Antibody Detection Kit (cPass; Genscript, Piscataway, NJ) uses a blocking ELISA format with human ACE-2 receptor molecules coated on an ELISA plate (12, 13). Human sera pre-incubated with labelled epitopes of the RBD on S1 proteins are then transferred to the plate. This blocking ELISA serves as a surrogate assay to determine the capacity of human sera to block the interaction between the Spike fusion protein (through its RBD) and its cellular receptor ACE-2. The analyte detected is anti-RBD blocking Ab of all subclasses. All the specimens, including positive and negative controls provided with the kit, were processed according to the manufacturer’s instructions, and included a 10X dilution factor of the primary specimen. All specimens and controls were tested in duplicate and the percentage of inhibition calculation was based on the mean of OD for each duplicate. A cut-off of 30% inhibition for SARS-CoV-2 neutralizing antibody detection was used to determine the presence of neutralizing antibodies, based on the manufacturer’s instructions for use.

### Abbott SARS-CoV-2 IgG assay

The Abbott SARS-CoV-2 IgG assay (Abbott Laboratories, Abbott Park, Illinois, USA), which detects IgG against SARS-CoV-2 N-protein, was performed on the Architect i2000sr platform according to the manufacturer’s instructions. Specimens were thawed on the day of testing and were centrifuged 10,000g for 10 min prior to each run. A sample to stored calibrator index (S/C) cut-off value of 1.4 was used for positive results, according to the manufacturer’s recommendations.

### STANDARD Q COVID-19 IgM/IgG Combo Rapid Test

The STANDARD Q COVID-19 IgM/IgG Combo Rapid Test (SD BioSensor, Gyeonggi-do, Republic of Korea) is a rapid immunochromatography test for the qualitative detection of specific IgM and IgG against SARS-CoV-2 N-protein on two separate test lines. Serum specimens were processed according to the manufacturer’s instructions. Briefly, 10μl of serum were applied to the specimen well of the test device. Three drops (90μl) of buffer were added immediately and vertically into the same specimen well. The test results were read visually at within 15 minutes. Any visible band was considered a positive result. To facilitate analysis of positive test results, the intensity of test bands was further classified as: no signal (0), barely visible but present (1), low intensity (faint but definitively positive) (2), and medium to high intensity (3) (Figure 1).

**Figure 1.**
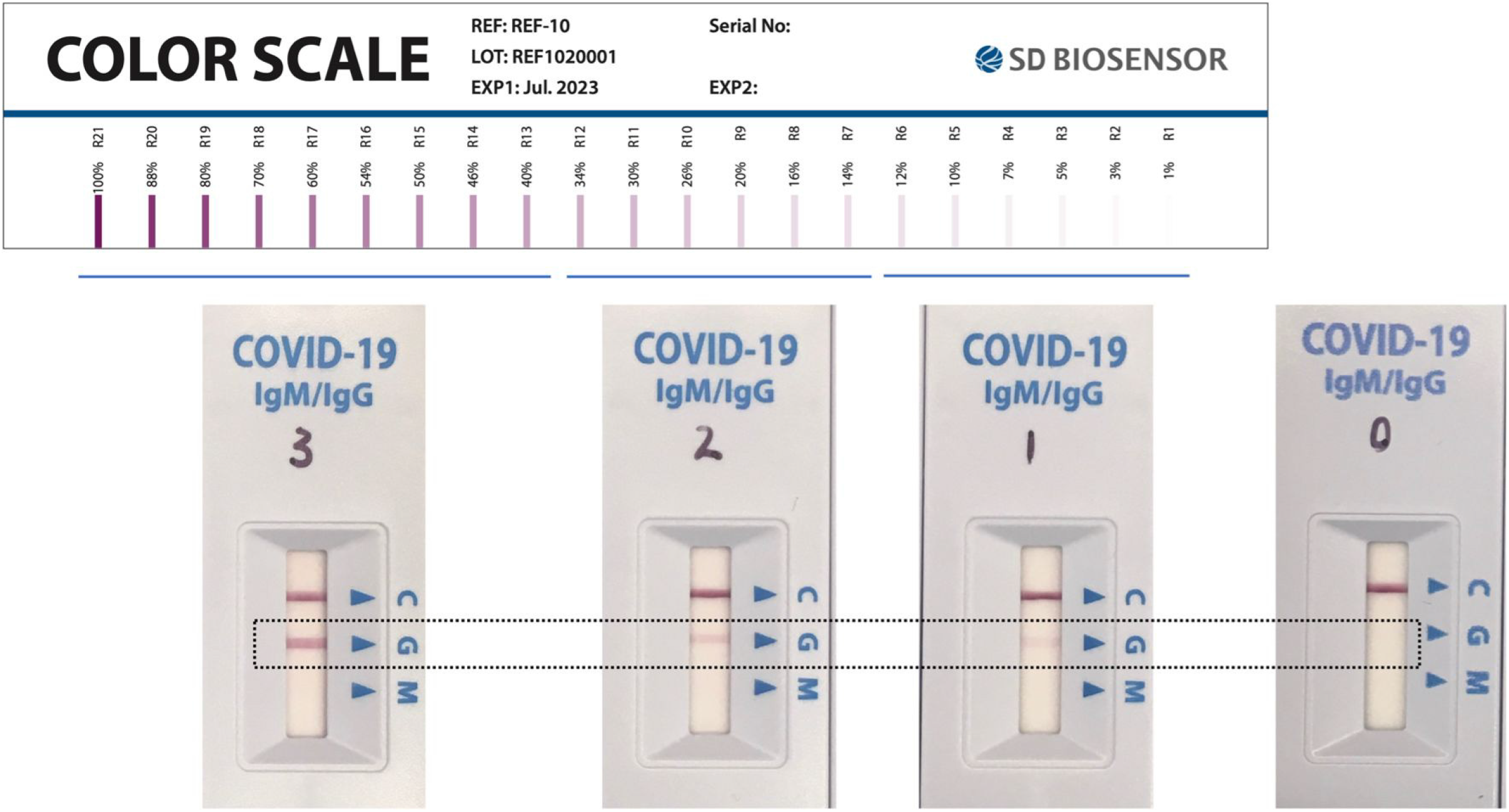
Categorization for SD RDT band intensity, based on a standard colour scale provided by SD Biosensor. A score of 0 indicates no signal; 1 indicates barely visible but present (corresponding to R1-R6 on the standard scale); 2 indicates low intensity (i.e. faint but definitively positive, corresponding to R7-R12 on the standard scale); 3 indicates medium to high intensity (corresponding to R13-R21 on the standard scale). The upper row shows the standard color scale provided by the manufacturer. The lower row shows actual RDTs used in the present study, photographed on the same day under standardized lighting conditions. The illustrative test line is shown in the dashed rectangle.

### Statistical analysis

Because all specimens were collected in the pre-pandemic era, prior to July 2019, all positive results for SARS-CoV-2 antibodies were considered false positives. The primary outcome calculated was test specificity and its corresponding 95% confidence intervals (95% CI), estimated according to a binomial distribution using Wilson Score method with Yate’s continuity correction as appropriate. The secondary outcome was relative risk (RR) for a false positive and the associated 95% CI. Both were estimated according to (i) positivity status for each parasite of interest, and (ii) SARS-CoV-2 target antigen tested. Statistical analyses were performed using R version 3.5.2 (R Core Team, Vienna, Austria). Area-proportional Venn diagrams were generated using eulerAPE version 3 (14).

## RESULTS

### Specificity of three commercial SARS-CoV-2 serological assays

The origin and characteristics of pre-COVID-19 specimens are reported in Table 1. Table 2 presents test specificity across the 559 samples tested. Overall, the point estimates of specificity of the cPass (10/559: 98%; 95% CI 97-99) and SD RDT IgG result (15/559: 97%; 95% CI 96-98) were similar to those for Abbott Architect (26/548: 95%; 95% CI 93-97) and SD RDT IgM result (30/559: 95%; 95% CI 92-96).

**Table 2.**
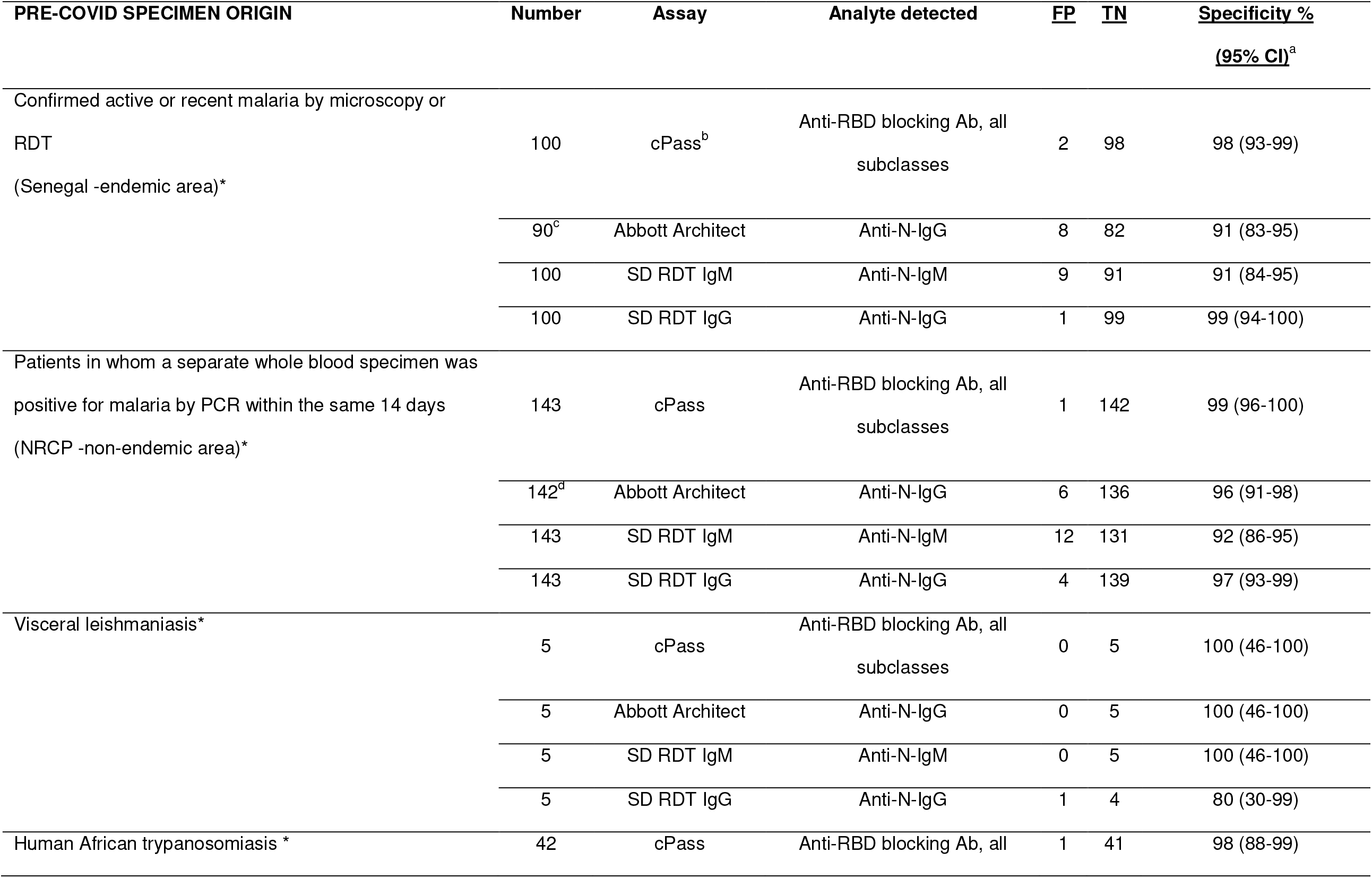
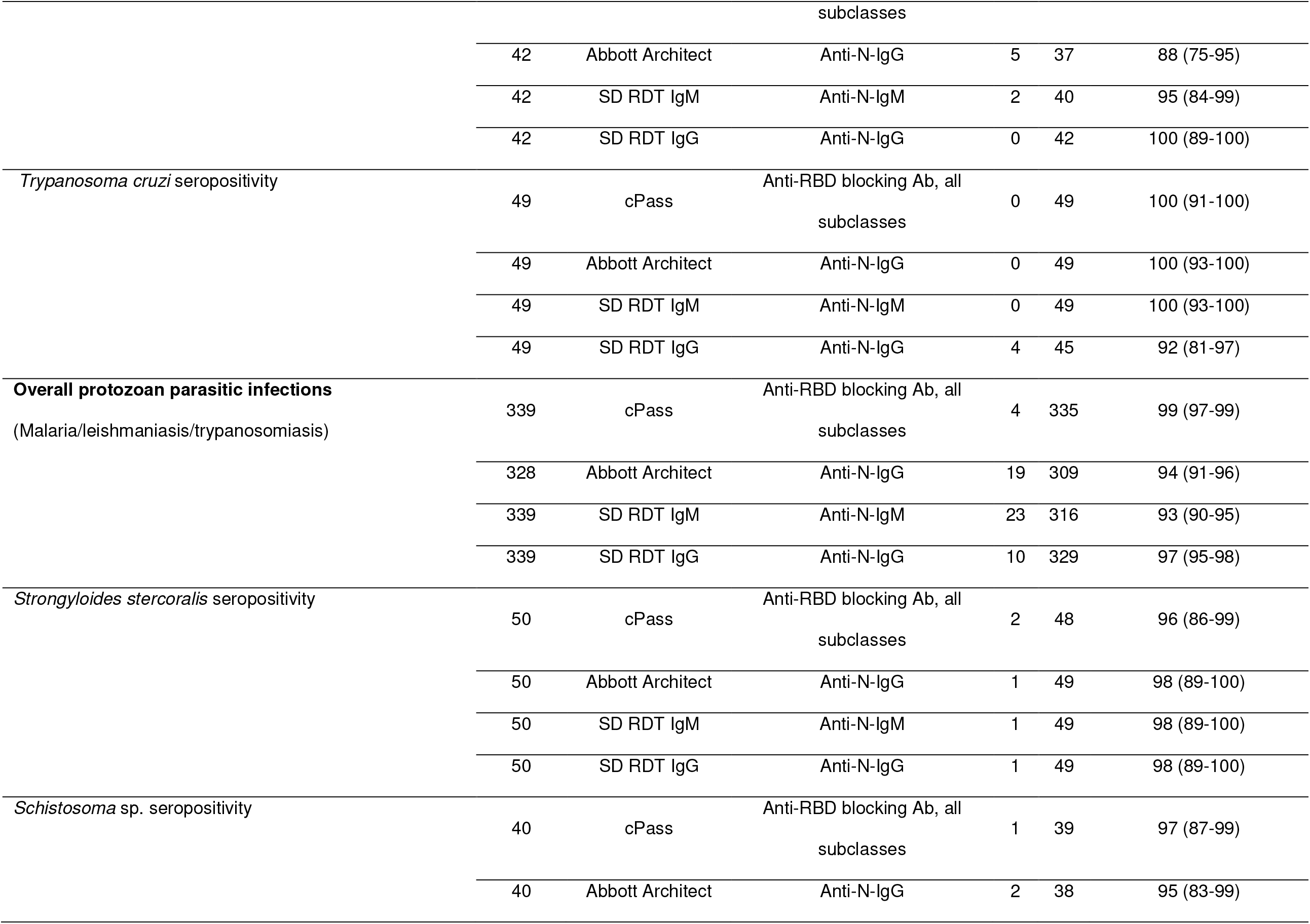
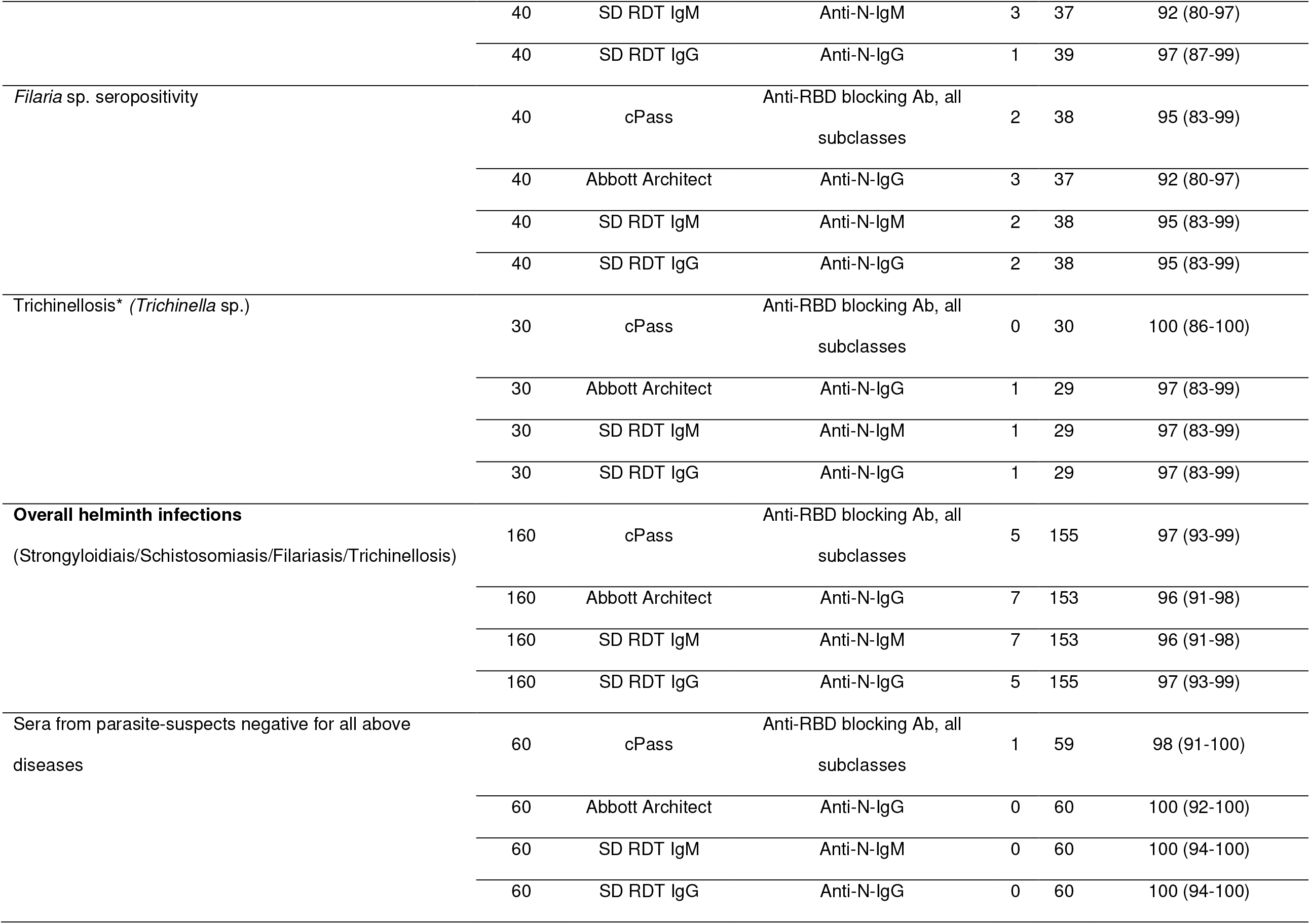
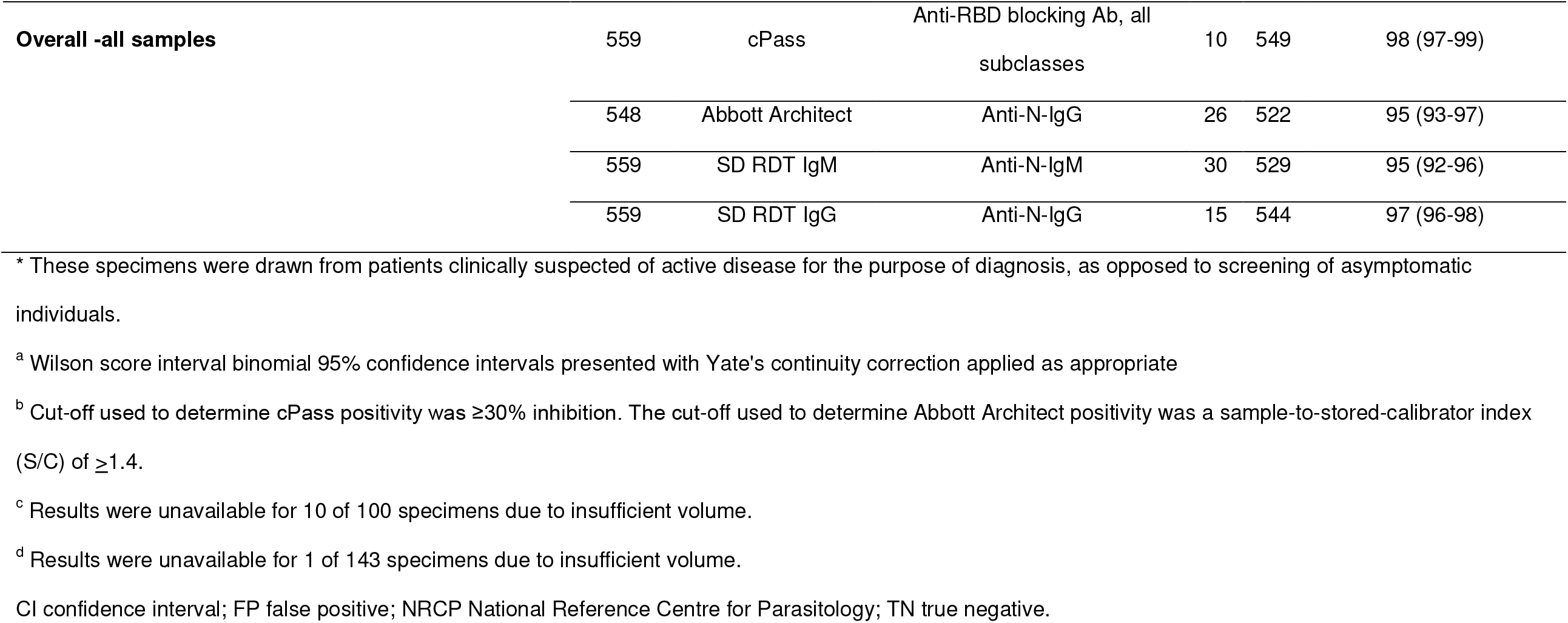
Diagnostic specificity of three commercial serological assays for the detection of SARS-CoV-2.

For specimens from patients with evidence of blood- or tissue-invasive protozoan infections overall, test specificity was as follows: cPass (4/339: 99%; 95% CI 97-99), SD RDT IgG results (10/339: 97%; 95% CI 95-98), Abbott Architect (19/328: 94%; 95% CI 91-96) and SD RDT IgM result (23/339: 93%; 95% CI 90-95). For specimens from Senegalese patients with malaria, specificity ranged from 91% (95% CI 84-95) for the Abbott Architect and SD RDT IgM result to 99% (95 % CI 94-100) for the SD RDT IgG result. For specimens from travellers with imported malaria, test specificity ranged between 92% and 99%. Among sera positive for anti-*Leishmania* sp and anti-*Trypanosoma cruzi* antibodies, cPass, Abbott Architect and SD RDT IgM displayed 100% specificity. However, the SD RDT IgG result yielded specificities of 80% (95% CI 30-99) and 92% (95% CI 81-97) against visceral leishmaniasis and human American trypanosomiasis, respectively. The SD RDT IgG result did not generate any false positives when used against human African trypanosomiasis specimens, whereas specificity of 88% (95% CI 75-95) was observed for Abbott Architect.

For specimens from patients with evidence of past or present helminth infection overall, test specificity estimates were all ≥ 96%. When evaluated against specimens seropositive for *Strongyloides* sp. (n=50) and *Trichinella* sp. (n=30), specificities ranged from 96% to 98%. Test specificity assessed against specimens seropositive for filarial species (n=40) ranged from 92% to 95%, and from 92% to 97% against specimens seropositive for *Schistosoma* sp. (n=40).

Sera collected from patients clinically suspected of parasitic infections that tested negative were also assessed. cPass yielded a specificity of 98% (1 false positive out of 60), whereas Abbott Architect, SD RDT IgG and SD RDT IgM showed a specificity of 100%.

To allow statistical comparisons across the entire group, we computed the relative risk (RR) and 95% CI of a false positive result by assay and target analyte, according to specimen origin (Table 3). Compared to cPass, the risk of a false positive SARS-CoV-2 result was higher for the Architect and the SD RDT IgM result overall across all specimens (RR 2.65, 95% CI 1.29-5.45; and RR 3.00, 95% CI 1.48-6.08, respectively), for malaria specimens overall (RR 4.89, 95% CI 1.42-16.79; and RR 7.00, 95% CI 2.11-23.16), and for protozoan infections overall (RR 4.91, 95% CI 1.69-14.28; RR 5.75, 95% CI 2.01-16.45). No significant differences were seen across assays for helminthic infections. SD RDT IgG relative risk of false positive was not significantly different to that of cPass for any of the specimen origins.

**Table 3.**
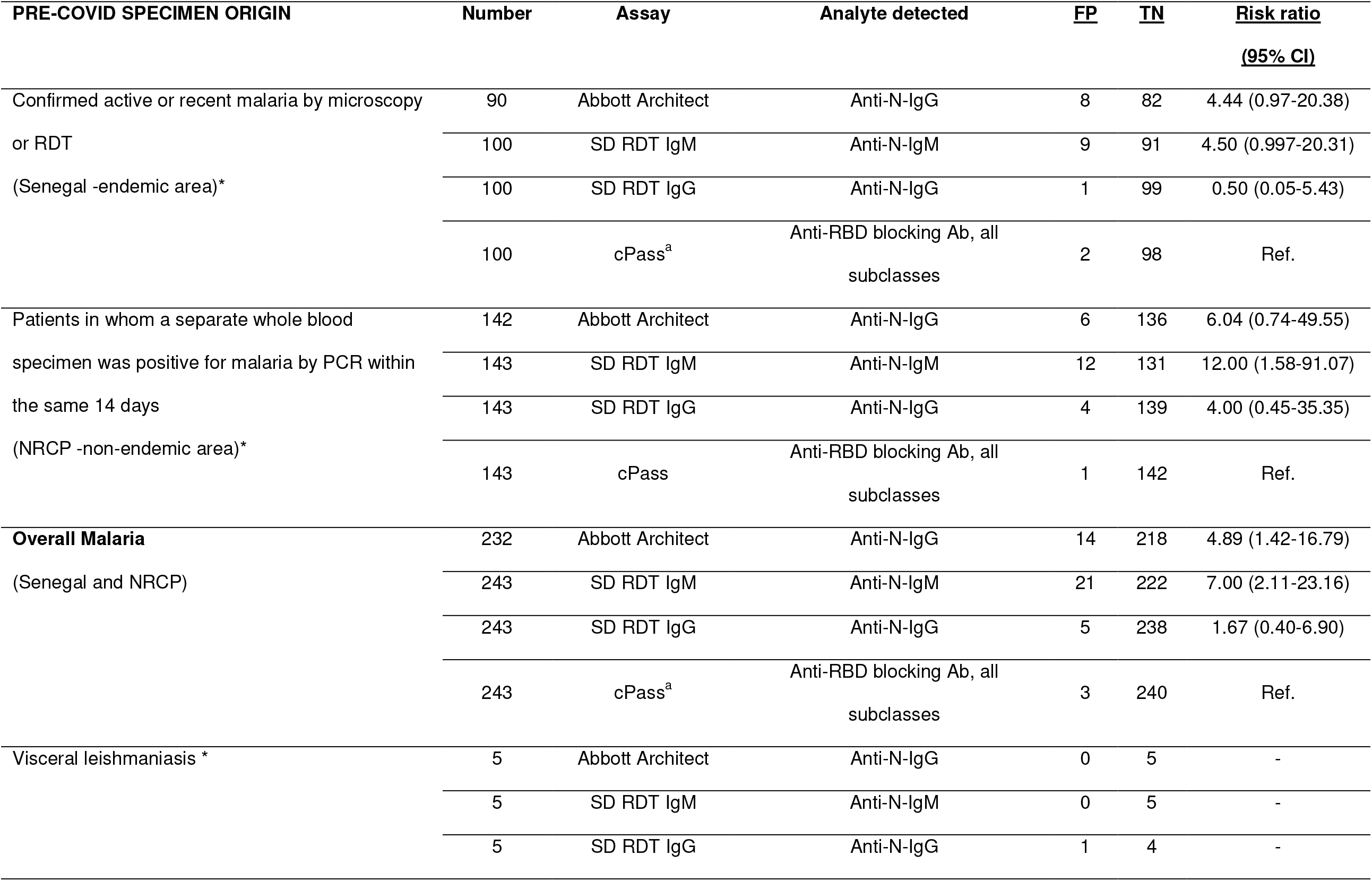
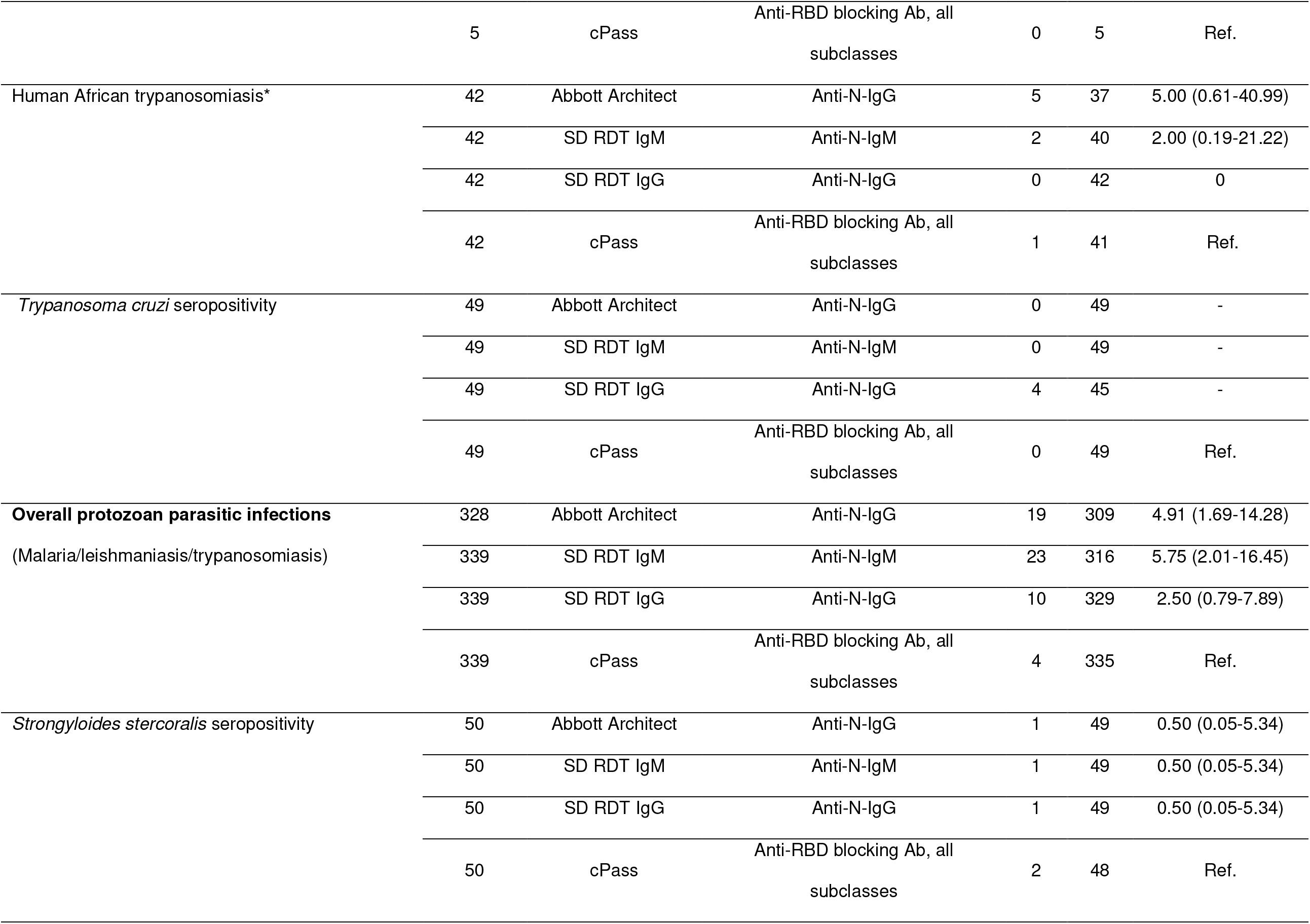
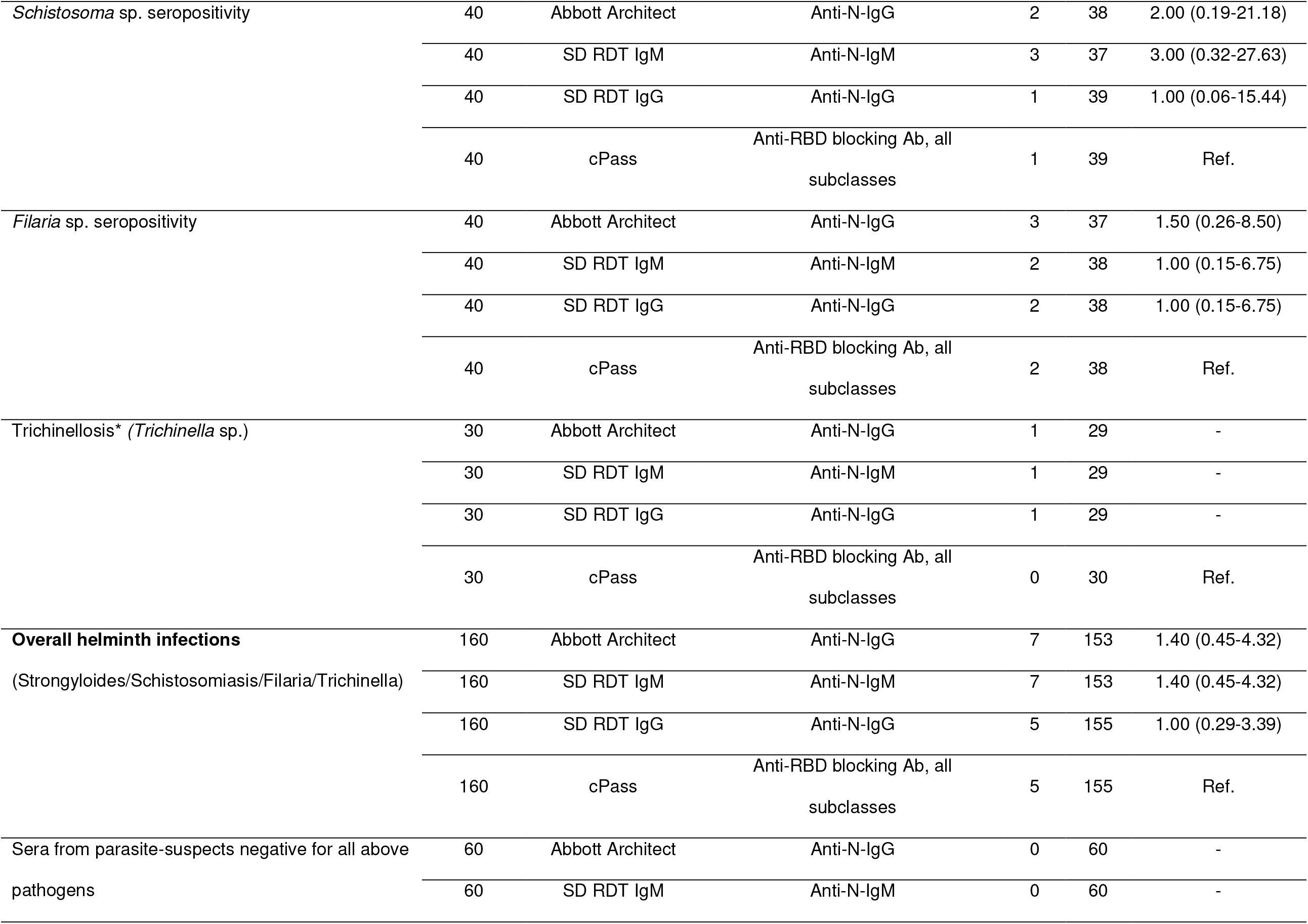
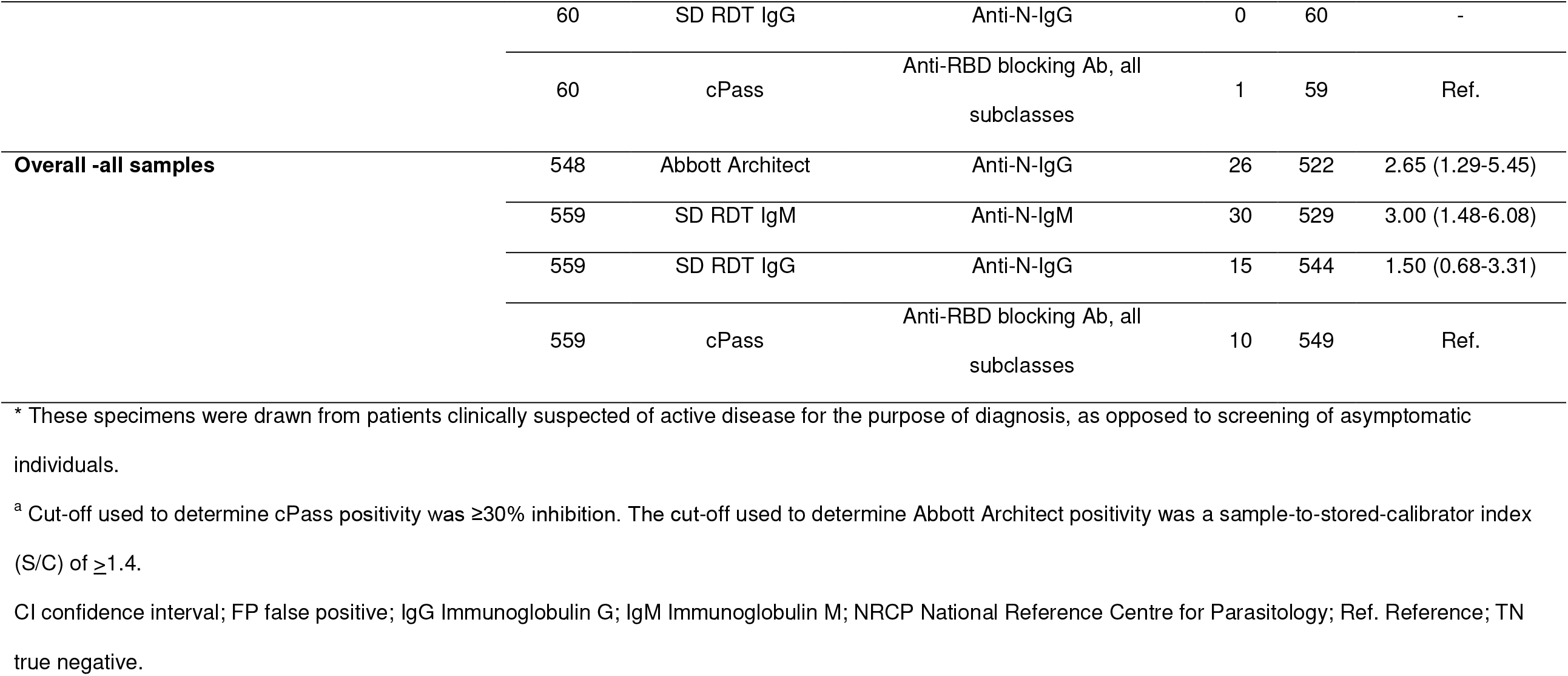
Relative risk of a false positive result by assay and target analyte, according to specimen origin.

### Characterisation of false positive results in terms of categorical agreement and signal intensity across serological assays

Categorical agreement between commercial serological assays for the detection of SARS-CoV-2 is depicted in Figure 2. The majority (>85%) of false positive results were positive by only one of the assays tested. When comparing cPass, Abbott Architect and SD RDT IgG (Fig. 2 A), cPass, Abbott Architect and SD RDT IgM (Fig. 2 B) or cPass, Abbott Architect and SD RDT IgG or SD RDT IgM (Fig. 2 C), all three assays were in agreement for only 14% (5/36), 7.8% (4/51) or 8.3% (5/60) of the false positive results, respectively. When comparing SD RDT IgG and SD RDT IgM (Fig. 2 D), the two assays were in agreement for 15% (4/39) of the specimens with a false positive result.

**Figure 2.**
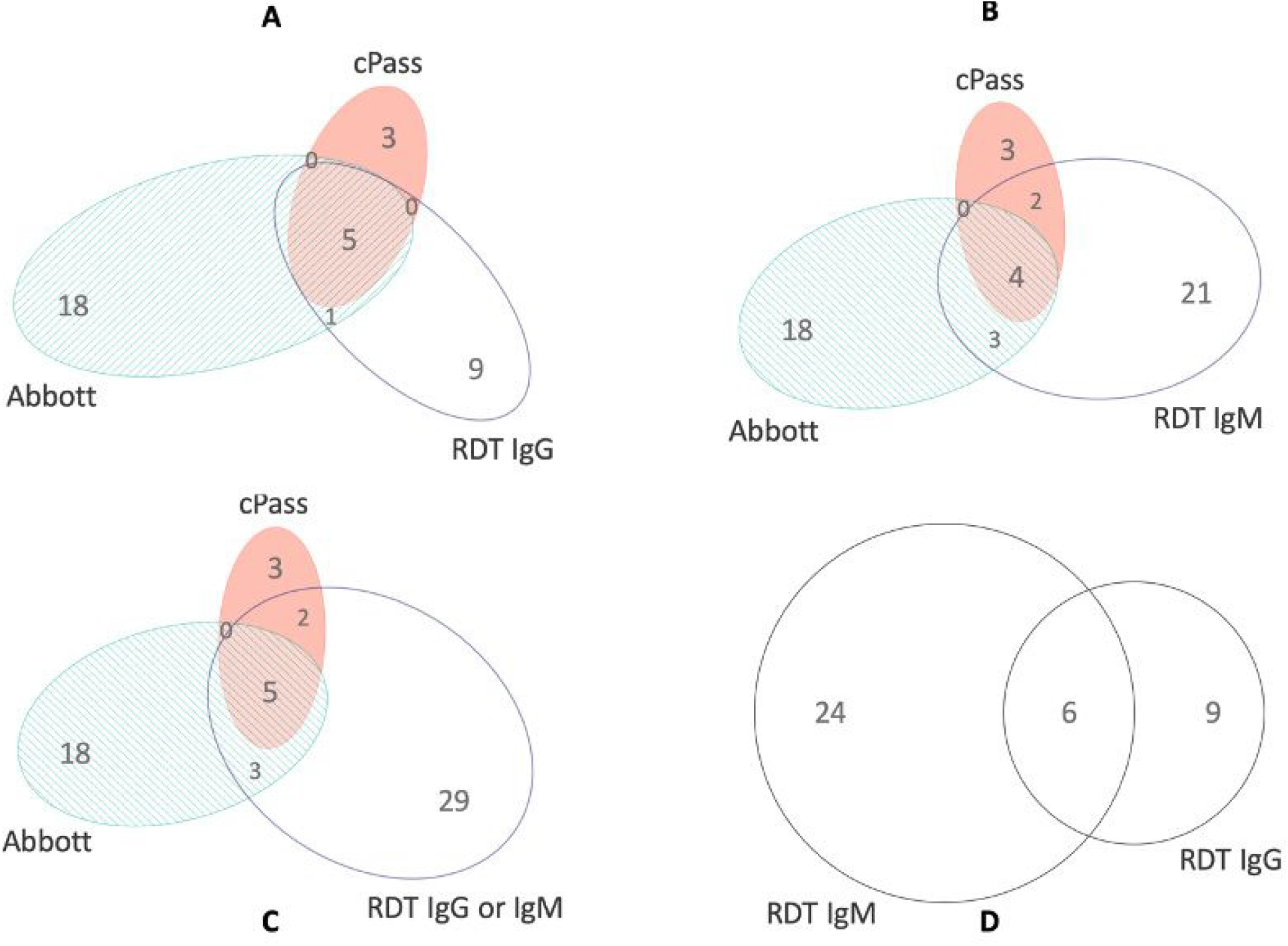
Venn diagram comparing false positive results from cPass, Abbott Architect, SD RDT IgG and SD RDT IgM serology from patients with protozoan and helminth parasites infections. Panels A, B, and C represent the overlap of cPass and Abbott Architect with SD RDT IgG, SD RDT IgM, or any positive SD RDT result, respectively. Panel D depicts the overlap of SD RDT IgG and SD RDT IgM. RDT denotes rapid diagnostic test; numbers denote the number of false positive specimens in each category.

Readout intensities of each serological test were assembled in a heatmap for specimens with false positive results from one or more tests (Figure 3). Overall, among specimens with false positive results, there was very little correlation between the signal intensity of a false positive test result and the probability of a false positive with another assay. Strong signal intensities were common among false positive results from laboratory-based assays. The cPass yielded positive results for 10/60 (16.7%) specimens with false positive results from one or more tests, with 5 of these having a binding inhibition of >50%. The Abbott Architect yielded positive results for 26/60 (43.3%) false-positive specimens, with 17 of these having sample-to-stored-calibrator index (S/C) of >1.68, which we considered as strong positives. By contrast, weak or very weak signal intensity was common for the false-positive results observed with the SD RDT. SD RDT IgG was positive among 16/60 (36.7%) false-positive specimens, with 5 of these having “barely visible but present” bands. By contrast, SD RDT IgM was positive for 30/60 (50%) false positive specimens, with 20 of these having “barely visible but present” bands. All but one of the other positive SD RDT IgM results were considered “weak”.

**Figure 3.**
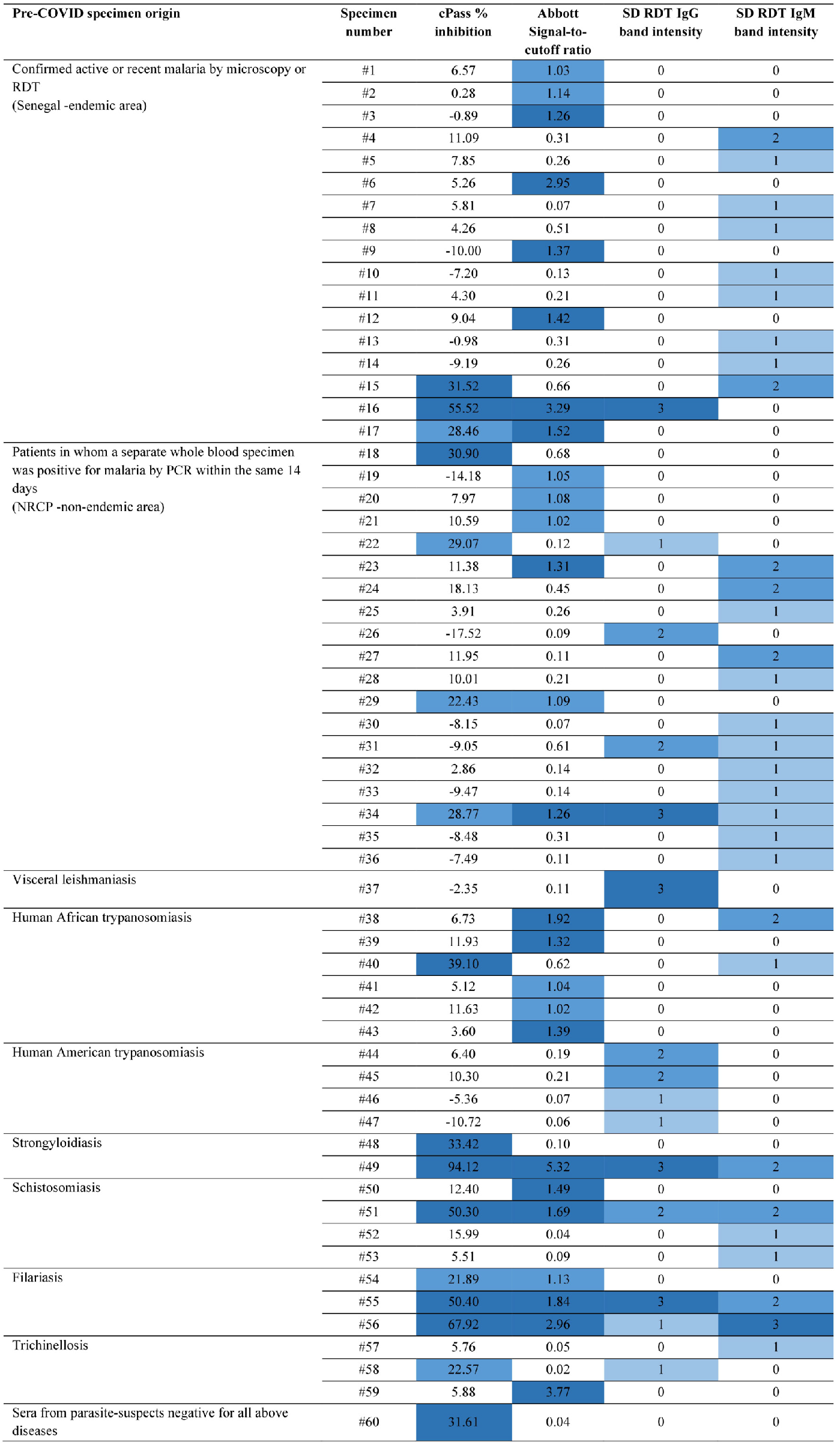
Heatmap of readout signal intensity of all false positive specimens identified using three commercial serological assays for the detection of SARS-CoV-2. RDT denotes rapid diagnostic test. Cut-off used to determine cPass positivity: Negative was <20% inhibition; Indeterminate was 20 to <30% inhibition; Positive was ≥30% inhibition. Cut-off used to determine Abbott Architect positivity: Negative was a signal-to-cut-off ratio <1.0; Weak positive was 1.0 to 1.2; Strong positive was >1.2 Categorization for SD RDT band intensity: a score of 0 indicates no signal; 1 indicates barely visible but present; 2 indicates low intensity (i.e., faint but definitively positive); 3 indicates medium to high intensity.

## DISCUSSION

We sought to assess the specificity of three SARS-CoV-2 antibody detection assays among people either with known tissue-borne parasitic infections or residing in areas where such infections are highly endemic. We tested assays against either the S or the N protein, among a large collection of well characterised pre-COVID-19 sera from clinical suspects with relevant tropical infectious diseases which may lead to cross-reactions with SARS-CoV-2 serologic assays. Previous reports found increased frequency of non-specific binding leading to positive results in smaller cohorts of patients with and without recent malaria in Nigeria (15), Benin (16) and Tanzania and Zambia (17). We confirm these findings with different serological assays and extend them to patients with imported malaria residing in a non-endemic area, as well as to patients with several key tropical infectious diseases for which there is a current void of available information on which to base interpretation of serological results for SARS-CoV-2.

The observed specificity of all assays ranged from 95-98% in the overall group of specimens. However, those for the Abbott Architect (95% [95%CI 93-97]) and the SD RDT IgM (95% [95%CI 93-96]) fell short of the WHO-recommended 97% benchmark for SARS-CoV-2 serological assays (18). Moreover, these values are well below estimates for the Abbott Architect from previous data among specimens from high-income countries, including 99.6% reported by the manufacturer using a panel of pre-COVID-19 specimens or from patients positive for alternative respiratory pathogens (n = 1070) (19) and 99.6% reported in an independent evaluation of 1099 pre-COVID-19 specimens (20). Similarly, the values for the SD RDT IgM are lower than the 100% specificity reported in the FDA serology test evaluation report for the STANDARD Q COVID-19 IgM/IgG Combo Rapid (21).

As expected, the lowest observed specificities were seen among sera from patients with protozoan infections of the reticuloendothelial system, such as human African trypanosomiasis (Abbott Architect; 88% [95%CI 75-95]), visceral leishmaniasis (SD RDT IgG; 80% [95%CI 30-99]), and from patients with recent malaria from a holoendemic area of Senegal (ranging from 91% for Abbott Architect and SD RDT IgM to 98-99% for cPass and SD RDT IgG). Non-specific cross-reaction among patients in malaria endemic areas may be potentiated by co-infections rather than from malaria infections alone. Alternatively, repeated infections with *Plasmodium* species rather than co-infections with other organisms may lead to a greater cross-reactivity. This is consistent with the association between false positive SARS-CoV-2 results and the presence of anti-*Plasmodium* antibodies (15), as well as the relatively higher specificity observed in our cohort of patients with antibodies to tissue-invasive helminths. Taken as a whole, the observed specificities among the assays and specimens tested are likely sufficiently elevated to be useful for serosurveys and epidemiologic tracking, but below the threshold required for individual patient care decisions (1, 18).

In order to allow statistical comparisons between different SARS-CoV-2 diagnostic assays, we computed the relative risk of a false positive result by diagnostic assay and target analyte, according to specimen origin (Table 3). The cPass showed the least variation across specimen origins and consistently had the highest specificity across groups. This assay was designed as a surrogate viral neutralization assay and measures the strength of inhibition of RDB binding to ACE-2 by host antibodies of any subclass. Perhaps surprisingly for a lateral flow immunochromatographic SARS-CoV-2 assay, the SD RDT IgG also showed very high performance across groups. Using cPass as the reference value, SD RDT IgG had a lower relative risk (RR) of a false positive result than either SD RDT IgM or Abbott Architect. The latter two tests were statistically significantly more likely to yield false positive results than the cPass for specimens with evidence of protozoan infections overall but not for specimens with evidence of tissue-invasive helminth infections. We previously showed that cPass has marginal advantages over anti-RBD IgG ELISA (12). In this case, the SD RDT IgG detects anti-N IgG and performed comparably to a sophisticated surrogate viral neutralization assay. Moreover, it compared favourably to the laboratory-based Abbott Architect in this group of specimens of interest. This is relevant to low-resource tropical settings where central laboratory capacity frequently limits care of patients with fever syndromes (22).

The finding of very low categorical agreement between SARS-CoV-2 serological assays among specimens with a false positive result is consistent with non-specific binding between host antibodies and test antigens. This observation can be leveraged to design testing algorithms with increased specificity. In our specimen set, requiring a positive result from a second test among cPass, Abbott Architect, or SD RDT IgG would rule out the majority of false positive results obtained after a first positive result (Figure 2). Others have proposed an avidity assay using various concentrations of urea washes to prevent non-specific binding (15) but this approach may not be suitable in low-resource settings, even when centralized laboratories exist.

Limitations of our study include the fact that the available volume of stored pre-pandemic specimens precluded the possibility of performing specific avidity testing or assessing for the presence of antibodies to seasonal coronaviruses that may have cross-reacted with the SARS-CoV-2 serological assays. However, a report from the United States found no false positives for Abbott Architect or SD RDT IgM/IgG among 21 patients with recent seasonal coronavirus infections: NL63 (n = 11), HKU1 (n = 7) and 229E (n = 3) (23). Moreover, the fact that our data recapitulates findings from previous studies in malarial-endemic areas is reassuring regarding their robustness.

## CONCLUSIONS

The specificity of SARS-CoV-2 serological assays was decreased among sera from patients with tissue-borne parasitic infections, below the threshold required for decisions about individual patient care. Specificity is markedly increased by the use of confirmatory testing with a second assay. Finally, the SD RDT IgG proved similarly specific to laboratory-based assays and provides an option in low-resource settings when detection of anti-SARS-CoV-2 IgG is indicated.

## Data Availability

After peer-reviewed publication, data available on request in the setting of a suitable research
protocol.

## FIGURE LEGENDS

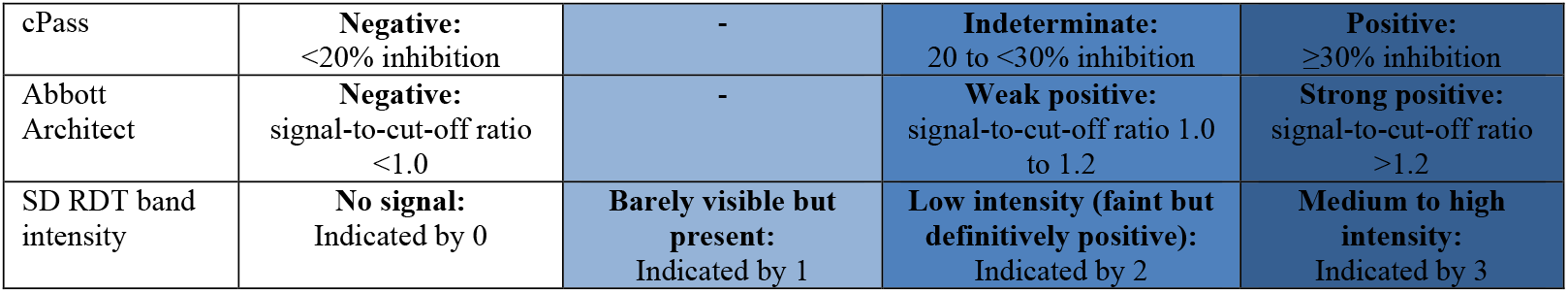

|  |  |  |  |  |
| --- | --- | --- | --- | --- |
| cPass | **Negative:**<br><20% inhibition | - | **Indeterminate:**<br>20 to <30% inhibition | **Positive:**<br>≥30% inhibition |
| Abbott<br>Architect | **Negative:**<br>signal-to-cut-off ratio<br><1.0 | - | **Weak positive:**<br>signal-to-cut-off ratio 1.0<br>to 1.2 | **Strong positive:**<br>signal-to-cut-off ratio<br>>1.2 |
| SD RDT band<br>intensity | **No signal:**<br>Indicated by 0 | **Barely visible but**<br>**present:**<br>Indicated by 1 | **Low intensity (faint but**<br>**definitively positive):**<br>Indicated by 2 | **Medium to high**<br>**intensity:**<br>Indicated by 3 |

